# Demographics, culture, and participatory nature of multi-marathoning – an observational study

**DOI:** 10.1101/2023.09.17.23295665

**Authors:** L Lundy, R.B. Reilly

## Abstract

**Objectives:** Completing 100 marathons defines a multi-marathoner.

The aim of this study was to document the sport of multi-marathoning and to report on the demographics, culture, and participatory nature of the sport.

**Methods:** A survey was distributed globally to the sports participants. All the major national and international multi-marathon clubs supported the survey distribution via their closed social media and email groups. The survey was fully anonymous and online.

**Results:** The survey attracted 830 respondents from 40 countries, with an average marathon completion count of 146 per respondent. 60% of respondents were men, with a respondent’s average age of over 50 years. Social life and travel superseded competitiveness as the main motivation as participants age. 57% of respondents had at least one contravention to a pre-participation screening questionnaire and 67% took pain relief medication around events. 93% of respondents reported that multi-marathoning was good for their mental health.

**Discussion:** Multi-marathoning facilitates the older athlete, but there is a significant gender imbalance in participation. Long-term health damage potential is significant, and health awareness requires addressing by governing bodies, event facilitators and participants alike. Multi-marathoners should use available information from modern equipment to inform their health status and to optimise training.

**Conclusion:** Recommendations from the study include implementing women-specific enhancements to events, race directors having adequately resourced health plans at events, and participants taking accountability for their own health by including proactive regular health management before and during their participation in the sport.

Summary Box

WHAT IS ALREADY KNOWN ON THIS TOPIC ⇒
Much research has been published on marathoning over recent decades, but none of the published studies focused on the sport of multi-marathoning.

WHAT THIS STUDY ADDS ⇒
Until now, absence of focused research has resulted in a lack of understanding of the demographics, culture, and participatory nature of the sport. This study is the first to provide this understanding.

HOW THIS STUDY MIGHT AFFECT RESEARCH, PRACTICE OR POLICY ⇒
Multi-marathon participants and those involved in multi-marathon governance, or in the running of multi-marathon events may utilise the results of this study to better plan their contribution to the sport and its overall safety, policies and organisation.

## Introduction

A boom in running in the 1970s and 1980s in the US and Europe focused on participation and jogging rather than competitive running [1]. After waning in the 1980s, it was revived in the late 1990s and continues to the present time [2, 3]. During this period, finishing organised events rather than just participation became an important aspect. It was during this increase in participation that multi-marathoning started. Multi-marathoner goals are based on the number of long-distance events completed rather than traditional faster/higher/stronger metrics. As there is less focus on performance in this sport, it is more inclusive, particularly for older athletes. Reaching the goal of 100 marathon completions defines a multi-marathoner.

A review of the literature showed much research has been published on marathoning over recent decades. ‘Marathoning’ has over 12000 references on the ‘PubMed’ and ‘Web of Science’ archives of biomedical and life science literature alone, but none of the published studies focused on the sport of muti-marathoning.

The main aim of this study was to address this lack of research on multi-marathoning, to report on the demographics, culture, and participatory nature of the sport and to document the sport. This was achieved via a globally distributed online survey that probed a wide range of facets of the sport. Embedded within the survey was the general health section of the international standard for vigorous sport pre-participation screening, the PAR-Q+ [4].

The study set out to address many and varied research questions that document the sport. These were:

- What is the age and gender profile of the multi-marathoner?
- What is the running history of participants in the sport?
- Are there any identifiable traits in terms of motivation?
- What goals do participants have?
- What is the health profile and what common injuries do participants commonly report?
- When injured, what treatment and recovery techniques do multi-marathoners follow?
- What running statistics are monitored and what are they used for?
- What is the diet profile of multi-marathoners?
- Do multi-marathoners use specific equipment?

It was hypothesised:

- multi-marathoning is a sport for older athletes and it is gender balanced.
- Participants have a long running history and value the social aspects of the sport as the primary motivation and achieving 100 marathon completions is their primary goal.
- Multi-marathoners are healthy and do not suffer from chronic injuries but do experience overuse injuries.
- Multi-marathoners are health aware and proactively engage medical resources when required.
- Multi-marathoners’ diet is varied, and they use the most advanced equipment available to them using available statistics to guide training plans.

## Methods

A survey was developed to engage with the multi-marathoning running community globally and address directly the study research. The survey was available on all common internet enabled devices and was optimised for mobile devices.

The survey was implemented using Qualtrics with a proxy server front end from Tiny URL giving a memorable survey custom URL https://multimarathon.study/survey [5].

The Trinity College Dublin’s Faculty of Health Sciences Research Ethics Committee (FREC) ethically approved the study recommending that the survey be distributed exclusively via gatekeepers with no direct approaches from the study team.

Gatekeepers were identified by analysing the structure of multi-marathoning at a national and international level.

All national and international multi-marathon clubs together with multi-marathon event companies in the UK and Ireland were approached by the study team and invited to act as gatekeepers for survey distribution to their members.

All the major national and international multi-marathon clubs proactively supported distribution of the survey through their social media channels or email groups. In addition, all the major UK and Ireland multi-marathon event companies similarly supported survey distribution to their members.

This approach ensured that the survey distribution was widespread and targeted specifically at the multi-marathon community.

The survey questions were wide ranging and were designed to document the demographics and the participatory nature in the sport of multi-marathoning and address the identified research questions relative to these demographics. 43 survey questions were grouped in the following areas:

- gender, age, and location.
- running history and performance.
- motivation including achievements and awards.
- diet.
- running equipment.
- running stats monitored.
- running injuries, recovery techniques and their efficacy.
- health:
  - participants physical readiness to participate in vigorous exercise based on PAR-Q+ [4].
  - pain relief medication while exercising [6, 7].
  - benefits on mental health and life stressors.
  - effects of SARS-CoV-2 on participants.

### Patient and Public Involvement

No patients or members of the public were involved in the design or interpretation of this study. All public participation was voluntary and informed consent was required before participation. We exclusively used gatekeepers for distribution of the survey to the target audience.

We intend to disseminate the main results to these gatekeepers for distribution.

### Equity, Diversity, and Inclusion

Our study included all genders, race/ethnicities, disabilities, and socioeconomic levels.

## Results

### Gender, Age and Location

The survey had 830 completed responses from 40 countries over 6 continents with a respondent average marathon completion count of 146.

Respondents had an average age of over 50 years with many participating in their 70s and 80s.

**Figure 1.**
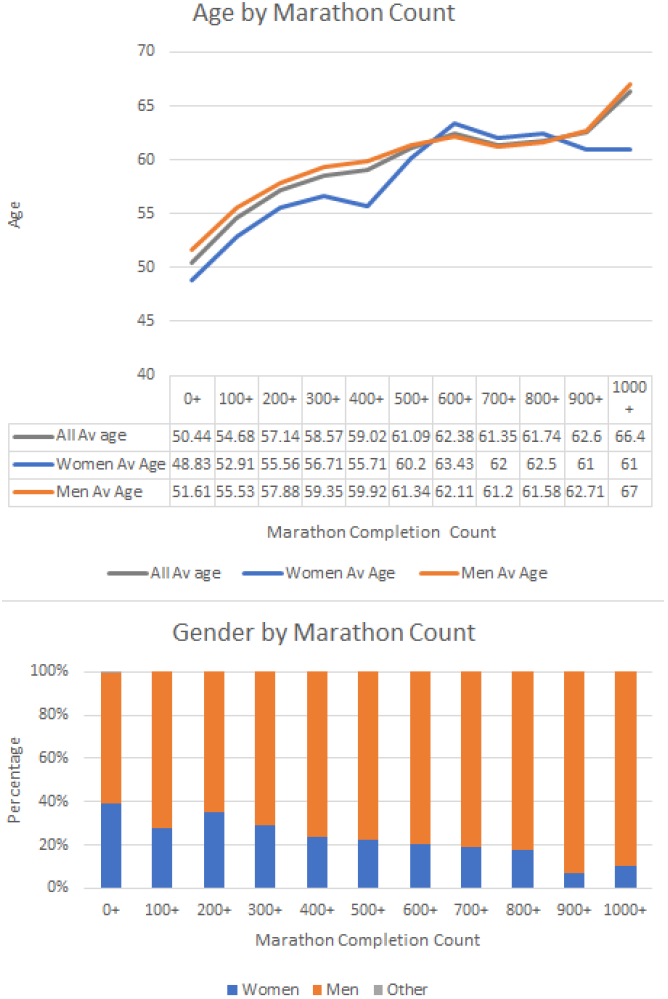
Age and Gender Distribution of Respondents

The survey demonstrated a significant gender imbalance with respondents being 60% men and 40% women.

The one notable exception to this gender imbalance was England, out of 269 respondents to the survey from England 44.6% were men and 55.4% were women.

### Running History and Performance

For those with over 100 marathons completions on average 80% complete at least one marathon a month and on average 12% complete at least one marathon a week.

**Figure 2.**
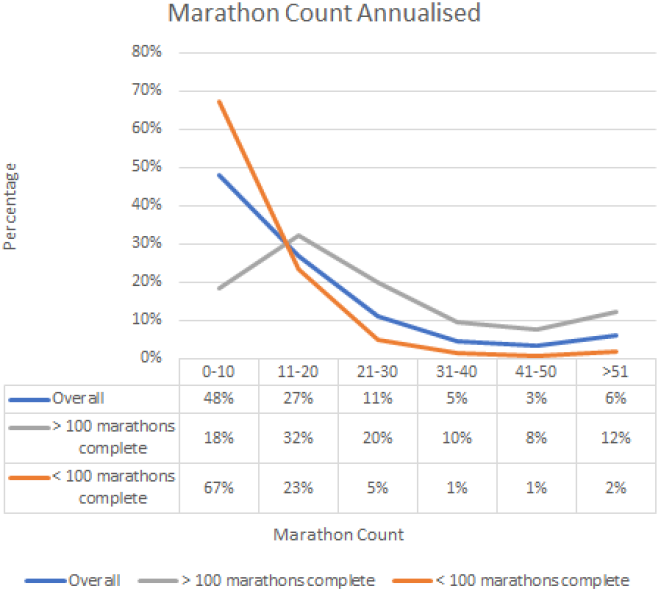
Survey Respondents Marathon Completion Count by annualised frequency

Performance of multi-marathoners was reported with most men having personal bests between 2:45-3:45 hrs., with a substantial number achieving less than 3:30 hrs. While most women reported having personal bests under 5:00 hrs., with a substantial number achieving less than 4:00 hrs.

Multi-marathoners reported that they run 57.91km a week including events. Effort was consistent throughout the year with many reporting that there was little peaking for specific events.

### Motivations, Awards and Achievements

Key reasons for participating in the sport were reported as:

- It was a way of life (13.63%)
- Keeping fit and healthy (18.32%)
- Sense of accomplishment (16.26%)
- Social life (11.53%)
- Reaching certain milestones (8.3%)
- Travel (8.82%)

Men with < 100 marathons completions reported being motivated by the need for continuous improvement and competition, but these motivations get replaced in the > 100 completions group where travel and social life take over as the main motivation.

Women prioritise social life, with reaching certain milestones being particularly important for women with < 100 marathon completions. Travel and social life were listed as more important in the women’s group with > 100 marathons completions.

Most multi-marathoners reported they were members of multi-marathon awards clubs, with 45% of respondents reporting achieving 100 marathon completions and gaining the 100-marathon completion award from their national club as being the biggest single goal.

### Diet

Most multi-marathoners claim that they do not follow a specific diet (69%). Of the other 31%, diet was important with significant numbers stating that they are vegetarian/ vegan/ paleo or pesco-vegetarian. Most of the non-categorised diets were vegetarian in nature.

**Figure 3.**
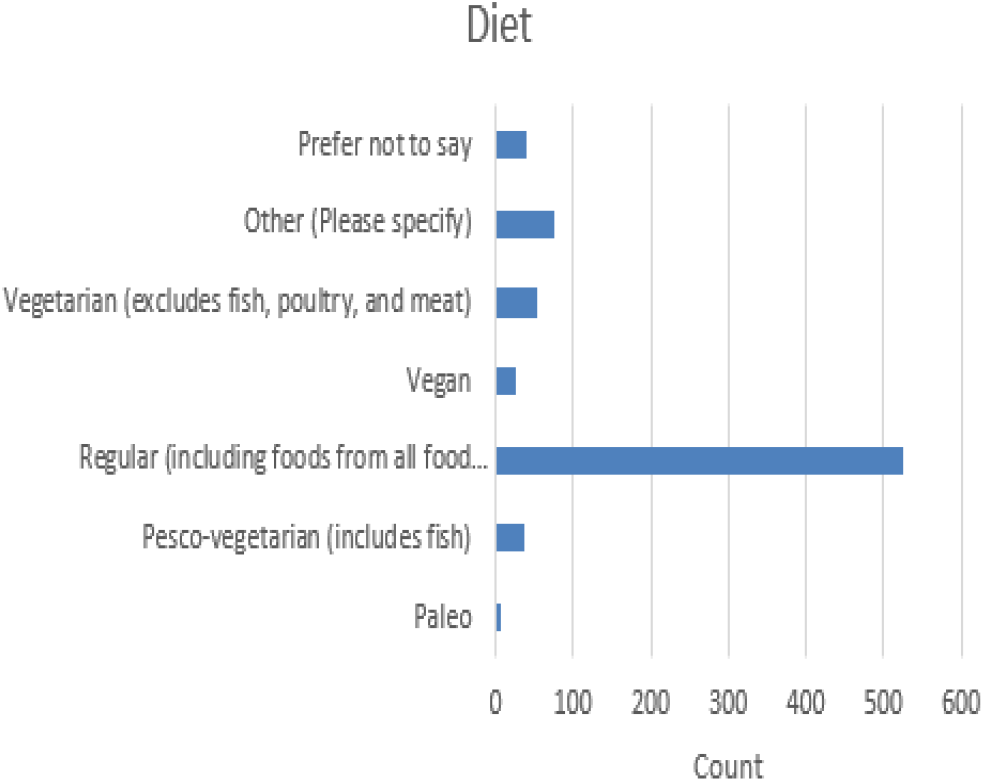
Survey respondents Diet Profile

### Running Statistics monitored

Steps, VO_2max_ and Heart Rate (HR) parameters were the most popular statistics monitored by multi-marathoners. In general, multi-marathoners monitor whatever stats were available on their wrist-based GPS device and this varies from device to device.

### Equipment

From the survey, running equipment was important to multi-marathoners with 93% reporting using GPS enabled devices. Through these devices wrist-based HR monitoring was commonplace but only 19% reported wearing the more accurate HR chest Strap. Only 22% said they use HR Zones despite this information being readily available [8-10].

44% of multi-marathons said they use headphones regularly for training (or events when allowed). With the introduction of bone conducting headphones, multi-marathoners have been at the forefront of their uptake with 41% of those who wear headphones reporting using this new headphone technology.

Cushioned/stability running shoes are the stalwart of the multi-marathoning community, with 45% of multi-marathoners reporting using cushioned shoes and 27% using stable shoes. 20% reported preferring light weight or minimal shoes.

**Figure 4.**
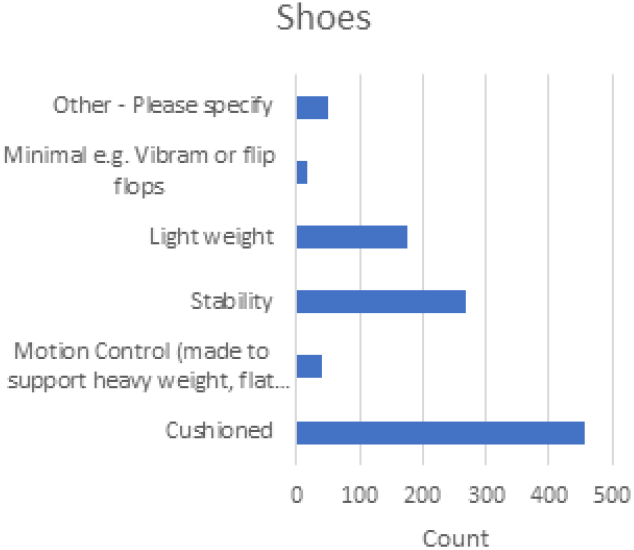
Shoe types used by Survey Respondents

### Injuries, Treatments and Recovery

The most common injuries reported were those associated with event-based issues blistering and chafing.[11]

Other injuries suffered over the running career were common running injuries which affected joints (ankle, knee, hip) and muscular injuries (calf, muscle pulls) or tendon issues (Achilles, IT Band). Plantar Fasciitis was also commonplace [12, 13]. No specific multi-marathoning injury types or specific overuse injuries were noted.

**Figure 5.**
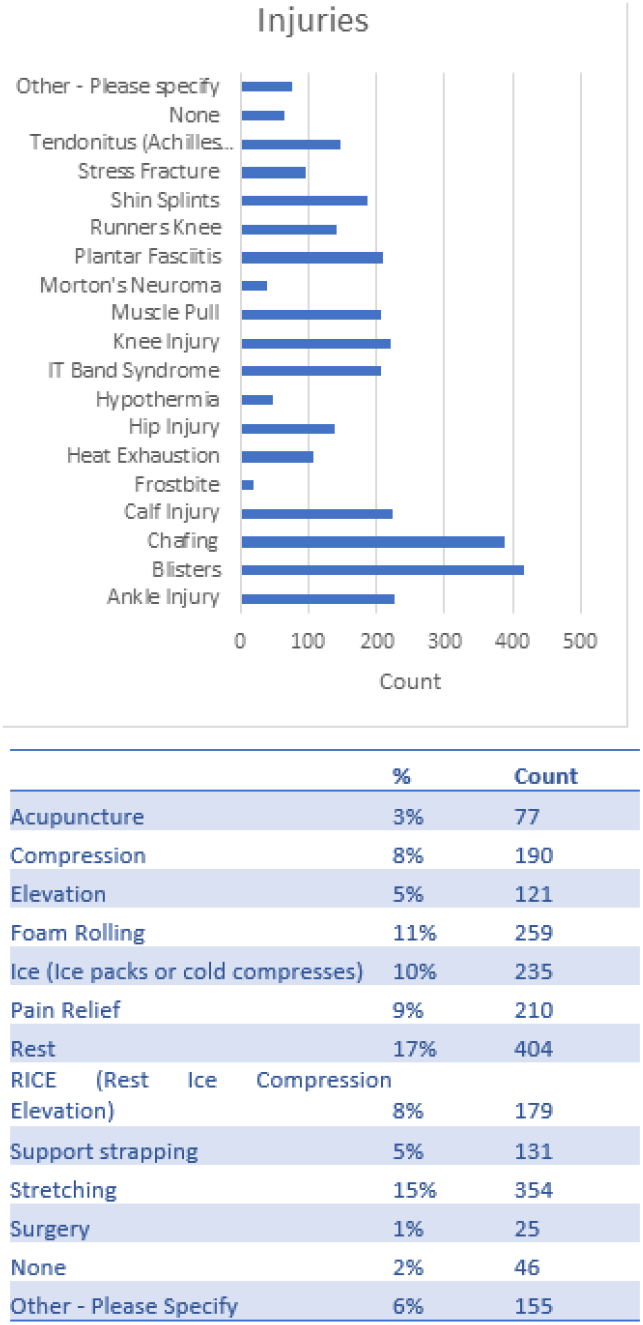
Injuries and Recovery Techniques reported by Survey Respondents.

When injured, 50% of multi-marathoners reported attending physiotherapists to manage the injury. 23% reported not seeking professional help at all.

Treatments vary widely with self-medication (stretching, rest, RICE, foam rolling and pain relief) being stated as the most popular.

### Pain Relief, Mental Health, and Life Stressors

Despite much information and discussion regarding the dangers of taking pain relief medication during extreme sport, 67% of respondents say they take medications that relieve pain around events [6, 7].

15% of respondents reported that their involvement in multi-marathoning had been a contributory factor to one of life’s stressors (e.g., relationship breakdown, long term illness, job loss).

More positively, 94% of respondents say that multi-marathoning was positive for their mental health.

### Health

The survey included the section on general health of the PAR-Q+ questionnaire, an internationally recognised vigorous event pre-participation screening questionnaire. Any contraventions of this section of the PAR-Q+ questionnaire require athletes to answer further questions, seek further information, or see a health professional before partaking in the sport [4].

Based on the survey 57% of respondents reported that they had at least 1 contravention while 25% had 2 or more contraventions to section 1 of PAR-Q+.

- 10% self-reported to have a heart condition.
- 13% self-reported to have a blood pressure issue.
- 9% self-reported to have had dizziness or lost consciousness in the previous year.
- 33% self-reported that they have bone, joint or tendon issues that would get worse with continued running.
- 17% self-reported to have other chronic medical conditions.

Of the respondents who self-reported having had chronic medical conditions (n=241), 47% were taking prescribed medication to control this condition.

71% of multi-marathoners had SARS-CoV-2 (Covid) by way of a positive test or thought that they had Covid. Many reported that symptoms persisted after eight weeks including: headaches, dizziness, tinnitus, septic arthritis, PoTS (Postural Tachycardia Syndrome), loss of taste and smell, excess perspiration, noise sensitivity, kidney pain, cough, respiratory inflammation, memory recall, asthma, arrhythmia, toe numbness, excessive blood pressure variation, low blood pressure.

**Figure 6.**
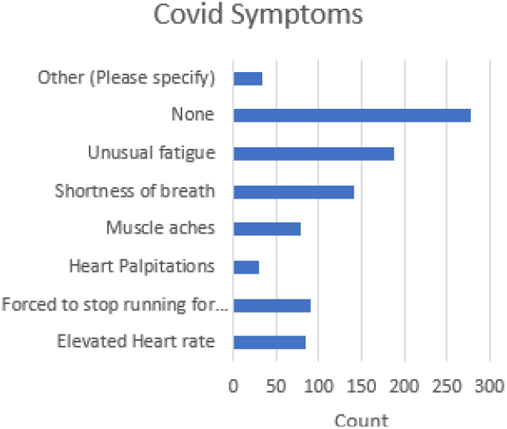
Reported Covid Symptoms

## Discussion

In line with the hypothesis, multi-marathoners are older athletes, and the survey results demonstrate that athletes can enjoy participation in the sport into their 70s and 80s. Due to the focus on the number of marathon events completed rather than performance, multi-marathoning has a particular attraction for older athletes.

This study recommends that participation in multi-marathoning should be encouraged as this enables older athletes to stay engaged with running and enjoy the health benefits of staying fitter longer [14].

Contrary to the hypotheses of gender equality, multi-marathoning is a male-dominated sport (M:60%, F:39%).

The one notable exception to this gender imbalance was England (M:45%, F:55%). The ‘England Model’ for events led to more even gender balance due to the maturity of dedicated multi-marathon race series, with:

- Unique 7-hour formats (e.g., traffic-free, no required navigation, short loops, enhanced social scene and flexible on-the-day distance choice).
- Specialist advisory group recommending women-specific enhancements for events.
- Many varied awards that encourage involvement.
- Vibrant social life around multi-marathoning.
- Popular award clubs and highly visible award t-shirts and ceremonies.

Together these actions encourage women to enter and to stay in the sport. This study recommends that the ‘England Model’ be adopted for events across other geographies.

In line with our hypothesis, the survey shows that reaching 100 marathon completions in the sport is the biggest goal. Achieving or working towards this goal defines a multi-marathoner. Typically, multi-marathoners take many years to reach this goal, but when achieved, multi-marathoning is considered by most to be a way of life. Indeed, for those multi-marathoners that had achieved this goal, 80% completed at least a marathon per month and 12% completed at least a marathon per week.

As hypothesised, social life is important to multi-marathoners and increasingly so as they age. However, other factors which vary by age and gender are also important. Men with less than 100 marathon completions tend to be motivated by the need for continuous improvement and competition. But this motivation gets replaced by those of travel and social life as they age, particularly after they achieve the goal of 100 marathon completions.

Women reported prioritising social life and reaching certain milestones from early in their involvement in the sport and, as with men, social life and travel get more important as they age. This study demonstrated that running multi-marathons can be enjoyed in later years, thus providing important social groupings and outlets for participants as motivations change.

Many survey respondents (31%) indicated that they follow a vegetarian style diet, with 5% vegan. To cater for this, multi-marathon race series increasingly offer vegetarian and vegan options at events. The study recommends that this should be encouraged as it caters for a significant sub-group in the multi-marathoning community.

Multi-marathoners were early adopters of running technology. They opt for stability and cushioning over lightness and performance when it comes to shoes, with less than 20% choosing performance-focused shoes.

Multi-marathoners prefer wrist-based devices with embedded HR monitoring capability. Only 19% reported that they regularly wear the more accurate chest-based HR monitors. Personalised information was readily available from these devices, and HR parameters are the most common health issue monitored. Contrary to our hypothesis, only a small number of multi-marathoners use HR Zones to aid training.

A recommendation arising from this study is the use of HR training zones to optimise training and set the correct training pace. It is also recommended to focus on HR parameter monitoring and heart health, in general, to minimise risk and maximise longevity in the sport.

Injuries within the sport of multi-marathoning tend to be normal running injuries and contrary to the hypothesis that multi-marathons predominantly suffer from overuse injuries, no specific overuse multi-marathon injuries or trends were noted [12, 13]. Temporary event-related issues were reported to be an ongoing concern of multi-marathoners (blisters, chafing) [11].

A substantial percentage (23%) of the respondents reported that they did not seek medical advice which is contrary to the hypothesis that multi-marathoners are health aware and pro-actively engage appropriate medical resources when required. A recommendation from his survey is that multi-marathoners seek professional medical help at the earliest opportunity during an injury cycle.

94% of respondents see multi-marathoning as good for their mental health, but 15% say that it was a contributary factor to life stressors (relationship breakdown, long term illness or loss of job). Though seen as a huge positive for mental health, further research is required to fully analyse the benefits.

Despite much information warning of the dangers of taking pain medication around extreme events, this study found 67% of respondents go against this advice and take medications that relieve pain. It is recommended by this study that multi-marathoners should avoid pain relief medication without medical advice and supervision [6, 7].

Contrary to our hypothesis that multi-marathoners are fit and healthy and do not suffer from chronic illnesses the survey demonstrated that significant numbers of multi-marathoners had contraventions to the general health section of the PAR-Q+ vigorous activity pre-screening questionnaire. This would require participants to answer further questions, seek further information, or see a health professional before partaking in the sport.

Multi-marathoners can be very cavalier with their own health despite being knowledgeable and having all the health statistics available to them. The study results indicate that all athletes on a multi-marathon journey should adhere to defined protocols before embarking on vigorous exercise (such as the PAR-Q+ questionnaire) and follow any recommendations provided. Further, they should also continue to have regular check-ups with medical professionals throughout their involvement in multi-marathoning and monitor all available health statistics for anomalies.

Most multi-marathoners reported contracting Covid (71%) with a large variety of symptoms, the most common being unusual fatigue and shortness of breath. Symptoms typically persisted beyond 8 weeks (Long Covid), with 10% forced to stop running for a period. Many reported having lingering symptoms long after contracting Covid. A follow-up study is required to understand the impact of Covid symptoms on multi-marathoners and assess pre-Covid/post-Covid participation and performance levels.

### Limitations

Those who responded to the survey are those motivated by multi-marathoning. Therefore, there is a selection bias among the respondents.

The survey was not able to capture comments from those who, for unknown reasons, abandoned the pursuit of this sport. The survey was only presented in English, which resulted in a smaller number of African and Asian respondents.

A follow-up study is required to better cover these regions.

### Research/Policy Implications

Until now, absence of focused research has resulted in a lack of understanding of the demographics, culture, and participatory nature of the sport. This study is the first to provide this understanding. Multi-marathon participants and those involved in multi-marathon governance, or in the provision of multi-marathon events may utilise the results of this study to better plan their contribution to the sport and its overall safety, policies, and organisation.

## Conclusion

This research seeks to inform and to guide those involved in the sport towards certain anomalies and recommends solutions to them. It highlights a significant gender imbalance and the propensity for a substantial number of multi-marathoners to participate despite reporting serious chronic illnesses.

These anomalies can be mitigated by implementing the ‘England model’ of events, race directors having properly resourced health plans at all multi-marathoning events and participants taking accountability for their own health by including pro-active regular health management before and during their participation in the sport.

## Abbreviations

FREC: Faculty of Health Sciences Research Ethics Committee
HR: Heart Rate
PAR-Q+: Physical Activity Readiness Questionnaire for Everyone

## Acknowledgements

The authors of this study would like to offer thanks to all study participants for their contribution to this research.

## Patient and public involvement

Patients and/or the public were not involved in the design, or conduct, or reporting, or dissemination plans of this research.

## Patient consent for publication

Not applicable.

## Provenance and peer review

Not commissioned, externally peer reviewed.

## Data availability statement

Data are available on reasonable request.

## Authors’ Contributions

Conception and design of all the research. LL, RBR All data acquisition and analysis and interpretation. LL, RBR, Drafting the work and revising it for important intellectual content: LL, RBR. All authors read and approved the final manuscript.

## Ethics Approval and Consent to Participate

Informed consent was obtained from all participants at the start of the survey and the study procedures were in accordance with the ethical requirements of the Faculty of Health Sciences Research Ethics Committee (FREC); Trinity College Dublin (TCD) which approved this study.

## Notes on Authors

**Leo Lundy**, BSc, is a PhD candidate at the School of Engineering at Trinity College, The University of Dublin.

**Richard Reilly**, PhD is Professor of Neural Engineering at Trinity College, The University of Dublin, a joint position between the School of Medicine and School of Engineering. He is Director and a principal investigator of the Trinity Centre for Bioengineering (TCBE) and also a principal investigator at the Trinity College Institute of Neuroscience (TCIN). His research focuses on high-density electrophysiological and neuroimaging-based analysis of sensory and cognitive processing for clinical applications.

